# Balance between maternal antiviral response and placental transfer of protection in gestational SARS-CoV-2 infection

**DOI:** 10.1101/2022.08.23.22279113

**Authors:** Juliana Gonçalves, Magda Melro, Marta Alenquer, Catarina Araújo, Júlia Castro-Neves, Nádia Charepe, Fátima Serrano, Carlos Pontinha, Maria João Amorim, Helena Soares

## Abstract

Maternal immune responses during pregnancy protect the growing fetus by clearing infection, preventing its vertical transmission, and through transplacental transfer of protective immune mediators to the fetus. How maternal immune response balances SARS-CoV-2 antiviral responses with transplacental transfer of protection to the fetus remains unclear. Our study shows that upon SARS-CoV-2 maternal infection, neutralizing antibodies (NAbs) are infrequently detected in cord blood. We uncovered that this is due to impaired IgG-NAbs placental transfer in symptomatic infection and to the predominance of maternal SARS-CoV-2 NAbs of the IgA and IgM isotypes, which are prevented from crossing the placenta. Crucially, the decision between favoring maternal antiviral response or transplacental transfer of immune protection to the fetus appears to hinge on the balance between IL-6 and IL-10 induced by SARS-CoV-2 infection, decreasing or increasing transplacental transfer of IgG-NAbs, respectively. In addition, IL-10 inversely correlates with maternal NK cell frequency. Finally, we found that ongoing infection favored perinatal transfer of maternal NK cells, highlighting a maternal sponsored mechanism to protect the newborn from horizontal transmission of infection. Our data point to an evolutionary trade-off which at once optimizes maternal viral clearance and vertical transfer of immune protection during the more susceptible perinatal period.

**Brief Summary:** In SARS-CoV-2 maternal infection, the balance between maternal antiviral response and transplacental transfer of cellular and humoral (NAb) protection hinges on maternal IL-6 and IL-10.

## Introduction

Maternal immune response protects the growing fetus by clearing the pathogen preventing its vertical transmission, and via transplacental transfer of protective cellular and humoral components to the fetus. This vertical transfer of protection continues after birth through secretory IgA and IgM antibodies and immune cells contained in breastmilk (1–3). Infections during gestation posit a challenge to the orchestration between priming of the maternal immune response in the tissue affected and the delivery of maternal immune components to the offspring, immediately via the placenta or posteriorly via breastmilk. The outcomes of SARS-CoV-2 infection in pregnancy vary from asymptomatic or mildly symptomatic to severe disease (4–6), with increased levels of inflammatory cytokines and immune cells being detected in maternal and cord blood samples (7, 8). How these cellular and humoral immune mediators balance maternal anti-SARS-CoV-2 response with the transplacental transfer of protection to the fetus remains to be addressed.

Maternal antibodies of the IgG isotype are transferred across the placenta from gestational week 13 through active transport mediated by FcRn expressed on synciciotrophoblasts (9). Due to their high molecular size and/or lack of specific transporters, maternal IgM and IgA antibodies are largely prevented from being transferred to the fetus (10). Neutralizing antibodies (NAb) targeting spike protein receptor-binding domain (RBD) have been shown to play an important role in controlling SARS-CoV-2 infection (11–13). Recent works in COVID-19 recovered individuals have put forward that SARS-CoV-2 neutralization is associated not only with anti-RBD IgG but to a significant extent to anti-RBD antibodies of IgA and IgM isotypes (14–17), raising the questions of how immune alterations during pregnancy impact the breadth of NAbs isotypes and consequently how efficiently they are transported across the placenta upon SARS-CoV-2 maternal infection. In addition to antibodies, immune cells and the cytokines they produce also play a role in resolution of SARS-CoV-2 infection, including CD4^+^ T (18) and NK cells (19). Namely, NK cells compose 20% of lung lymphocytes and play an important role in eliminating SARS-CoV-2 infected cells (19). Pregnancy leads to a decrease in CD4^+^ T and NK cell levels in circulation (20). Nevertheless, increased frequency of NK cells has been detected in neonates born to pregnant women with ongoing infection (8) and neonatal NK cells have been shown to display antiviral response from birth (21). In COVID-19 patients the profile of inflammatory cytokine responses has been associated with disease outcomes. While an interferon response is conducive to viral clearance (22), the enduring production of inflammatory cytokines IL-1b, IL-6, IL-8, IL-10 and IL-18 has been linked to disease severity (23, 24). Pregnant women have been described to mount a mild inflammatory response to SARS-CoV-2 infection that is reflected in the fetus, even in the absence of vertical viral transmission, as previously observed for gestational HIV-1 and HBV infections (7, 8, 25, 26).

In view of the potential long-term harm to the developing fetus it is important to identify how long cytokines linger even after the infection has been cleared, the extent that they are transferred to the fetus and to further document possible fetal immune priming (8). Crucially, it remains to be addressed whether there is a cellular component to SARS-CoV-2 transplacental transfer of immune protection and crucially, the identification of the inflammatory networks regulating the transfer of protective antibodies and immune cells from the infected mother to the fetus.

In this study, we advance a role for NK cells and anti-spike IgM in balancing maternal antiviral response with vertical transfer of immune protection. We identified that breadth of SARS-CoV-2 NAbs isotypes depended on gestational age at the time of infection. While second or earlier third trimester SARS-CoV-2 NAbs were exclusively associated with anti-spike antibodies of IgA and IgM isotypes, in ongoing infections NAbs correlated with anti-spike IgA, IgG and IgM. In addition, we show that in symptomatic ongoing infections IgG-NAbs transplacental transfer in hindered. Consequently, we demonstrate that the transfer of NAbs, but not of anti-spike IgG, to the fetus is impaired. The role of anti-spike IgM antibodies and NK cells in gestational protection from SARS-CoV-2 infection is further highlighted by their association with asymptomatic maternal infection. Mechanistically, we have put forward that the decision between favoring maternal antiviral response or transplacental transfer of immune protection to the fetus appears to hinge on the balance between IL-6, and IL-10. IL-6 and IL-10 are associated with decreased or increased IgG-NAbs transplacental transfer, respectively. In addition, IL-10 inversely correlates with the frequency of maternal NK cells, in circulation. By interpolating our experimental results with clinical data, we identified that ongoing infection favored the transfer of maternal NK cells to the neonate perinatally. Altogether our data support a two-pronged model, whereby maternal immune responses might protect the neonate from both vertical and horizontal SARS-CoV-2 transmission. First, through early NK cell and IgM mediated viral clearance, preventing in utero viral transmission. Second, by favoring a maternal sponsored mechanism to protect the newborn from horizontal transmission of infection, a time when close physical contacts between mother and neonate together with neonate’s immature immune system results in increased risk of infectivity. Our data support that this might be achieved not only via perinatal transfer of NK cells, but crucially through the transfer of SARS-CoV-2 NAbs of the IgA and IgM isotypes through breastmilk.

## Results

### Population

Our study comprised 72 pregnant women recruited from May 2020 to February 2021, before the introduction of COVID-19 vaccination to the general population. Sixty pregnant women tested positive (CoV-2^+^; Table 1) and 12 tested negative (CoV-2^-^; Table 2) for SARS-CoV-2 by PCR on nasopharyngeal swabs (Figure 1A). Of the 60 SARS-CoV-2^+^ pregnant women, 9 tested positive in the second trimester (recovered second trimester group, 2R; Figure 1A), 11 tested positive between 20 to 154 days before delivery (recovered third trimester group, 3R; Figure 1A), and 40 tested positive in late 3^rd^ trimester within 11 days of delivery (ongoing third trimester infection group, 3O; Figure 1A). Fifty percent of pregnant women were symptomatic, with four pregnant women being admitted to the hospital due to COVID-19 symptomatology, non-invasive oxygen support was provided to the other three. Within the SARS-CoV-2^+^ population, 50 mother-umbilical cord matched samples were collected at the time of birth, and in 82% of the deliveries a nasopharyngeal swab was performed to the newborn, with 3 babies testing positive but remained asymptomatic and one baby that tested negative being admitted to neonatal ICU (Table 1). Additional 9 pregnant women were recruited between July of 2021 and February of 2022 having undergone COVID-19 mRNA BNT162b2 vaccine. Maternal and cord blood samples were collected from 6 dyads (Figure 1A; Table 3).

**Figure 1.**
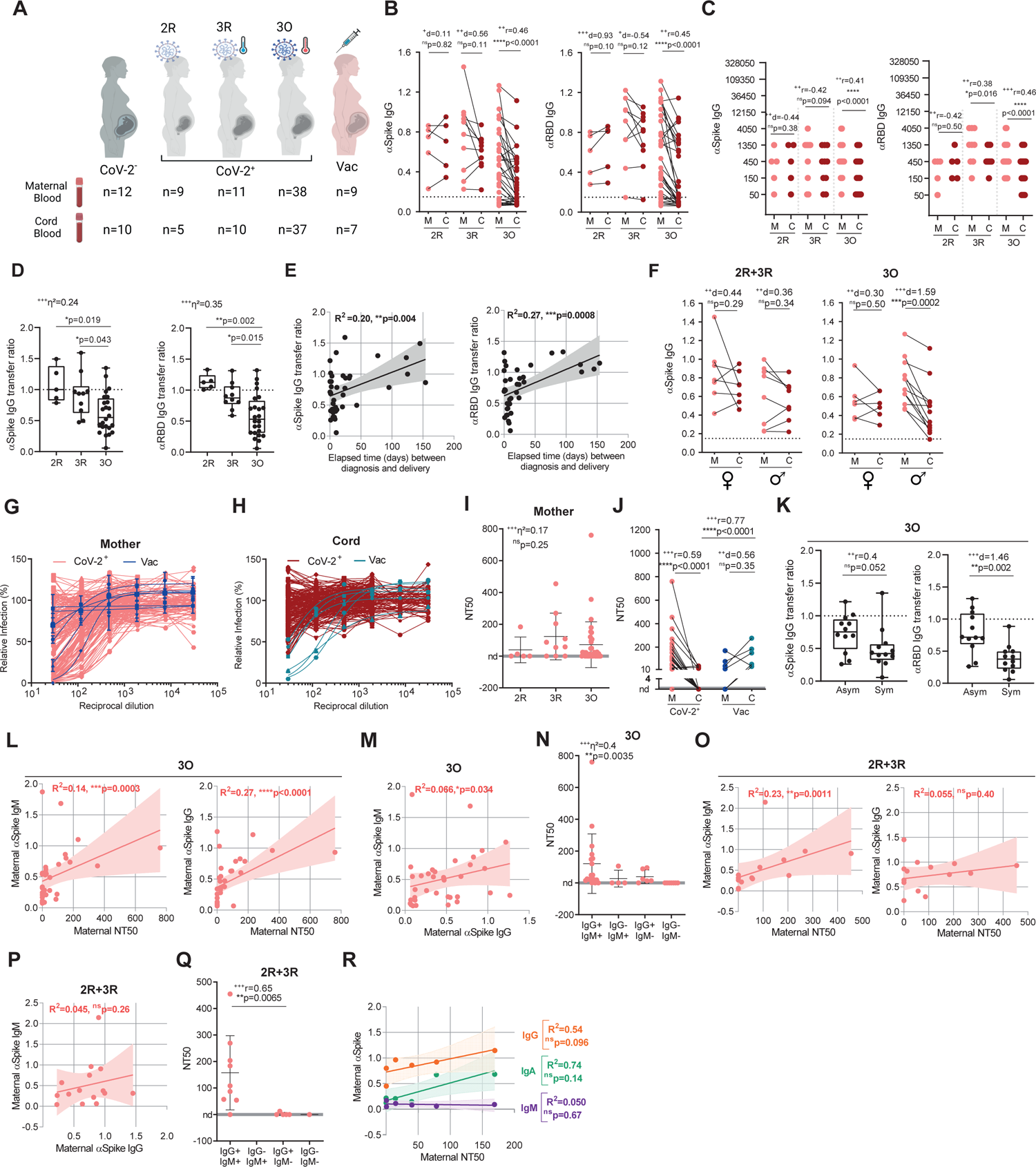
Low NAbs in neonates are associated with maternal production of IgM-NAbs and impaired transfer of IgG-NAbs in symptomatic infections. A-Outline of participants recruitment, divided in CoV-2^-^ group of pregnant women negative for SARS-CoV-2 (n = 12); in 2R group (recovered at delivery) of pregnant women that tested positive for SARS-CoV-2 in the second trimester (n = 9); in 3R group (recovered at delivery) of pregnant women that tested positive for SARS-CoV-2 earlier in the third trimester (n = 11); 3O group (ongoing infection at delivery) of pregnant women that tested positive for SARS-CoV-2 at the time of delivery (n = 38); Vac group includes pregnant women that were vaccinated with COVID-19 BNT162b2 vaccine (n = 9). B-Paired dyad analysis of anti-spike (left) and anti-RBD (right) IgG in 2R, 3R and 3O groups. Dashed line indicates assay cutoff (n = 50 dyads). C-Paired dyad endpoint titers for anti-spike (left) and anti-RBD (right) IgG in 2R, 3R and 3O groups (n = 50 dyads). D-Anti-spike (left) and anti-RBD (right) IgG transfer ratio in 2R, 3R and 3O groups (n = 39). E-Correlation between anti-spike (left) and anti-RBD (right) IgG transfer ratio and elapsed time between diagnosis and delivery (n = 39). F-Paired dyad analysis of anti-spike IgG levels in 2R+3R and 3O groups according to neonate sex (2R+3R n = 15 dyads; 3O n = 18 dyads). G-Neutralization curves of maternal plasma from SARS-CoV-2 positive (CoV-2^+^, n = 50) and vaccinated (Vac, n = 6) participants. H-Neutralization curves of cord plasma from SARS-CoV-2 positive (CoV-2^+^, n = 50) and vaccinated (Vac, n = 6) participants. I-Neutralization titer (NT50) of maternal plasma from SARS-CoV-2 positive individuals from 2R, 3R and 3O groups; nd: not detectable (n = 50). J-NT50 paired dyad analysis in SARS-CoV-2 positive (CoV-2^+^, n = 50) and vaccinated participants (Vac, n= 6). K-Anti-spike (left) and anti-RBD (right) IgG transfer ratio in 3O group in function of the presence (Sym) or absence (Asym) of symptoms (n = 24). L-Correlation between maternal neutralization titers (NT50) and anti-spike IgG (left) or anti-spike IgM (right) in 3O group (n = 35). M-Correlation between maternal anti-spike IgM and IgG levels in 3O group (n = 35). N-Maternal NT50 in function of the presence of anti-spike IgG and/or IgM antibodies in 3O group (n = 35). O-Correlation between maternal neutralization titers (NT50) and anti-spike IgG (left) or anti-spike IgM (right) in 2R+3R groups (n = 15). P-Correlation between maternal anti-spike IgM and IgG levels in 2R+3R groups (n = 15). Q-Maternal NT50 in function of the presence of anti-spike IgG and/or IgM antibodies in 2R+3R groups (n = 15) R-Correlation between maternal NT50 and anti-spike IgG (orange), IgA (green) and IgM (purple) in vaccinated group (n = 6). nd: not detectable; p values *p < 0.05, **p < 0.01, ***p <0.001, ****p < 0.0001; ns, not significant were determined by parametric paired t test (B, C, F, J), non-parametric paired Wilcoxon test (B, C, J), unpaired t test (K), Mann-Whitney test (J, K, Q), ordinary ANOVA, post hoc Holm-Sidak (D), Kruskal-Wallis test, post hoc Dunn’s (I, N), Pearson correlation (E, O) and Spearman correlation (L, M, P, R). Effect size measures ^+++^high, ^++^medium, ^+^small were determined by d = Cohen’s d (B, C, F, J, K), r = correlation coefficient r (B, C, J, I, K, M, Q) and η^2^ = eta-squared (D, I, N).

**Table 1:**
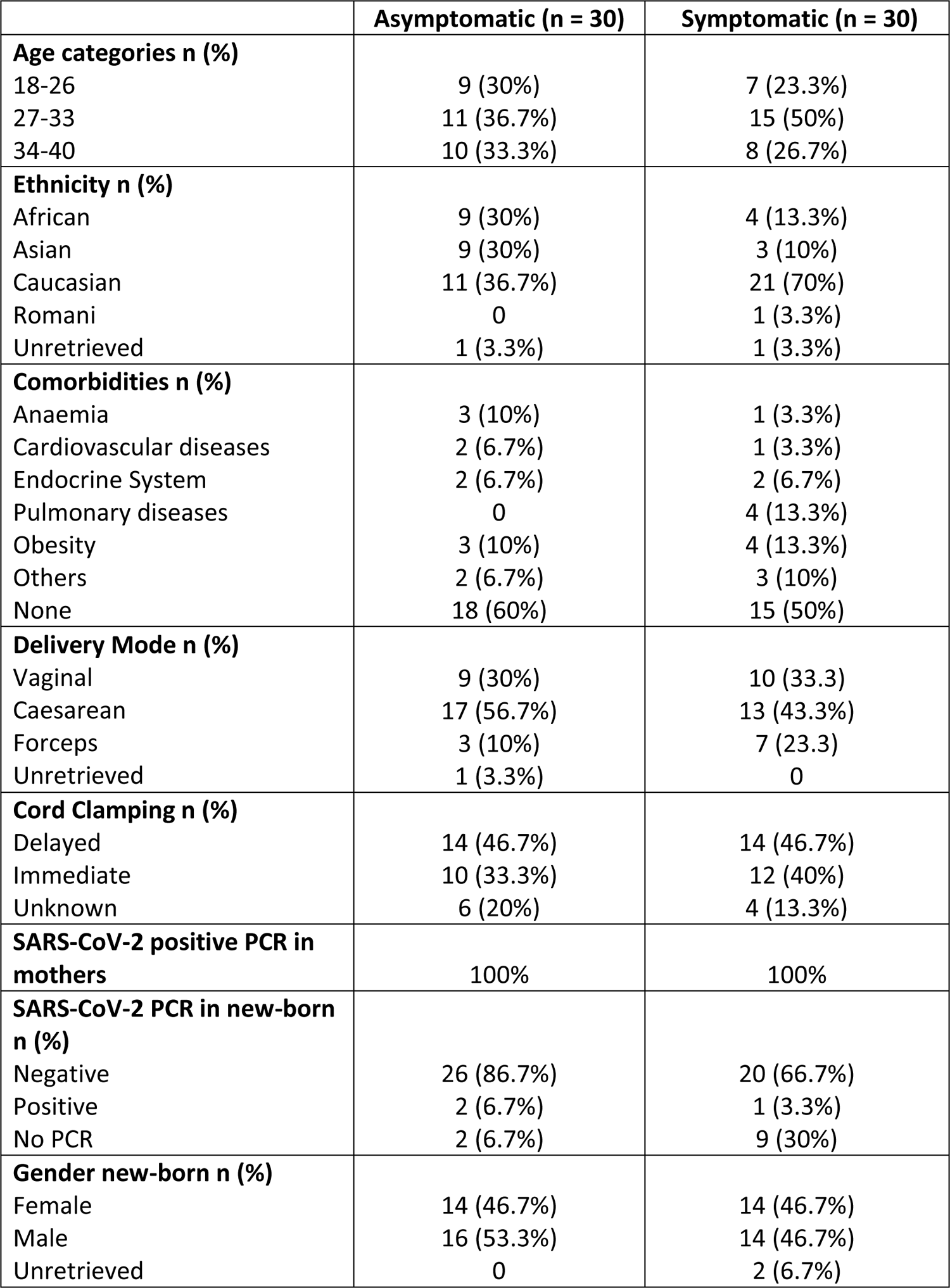
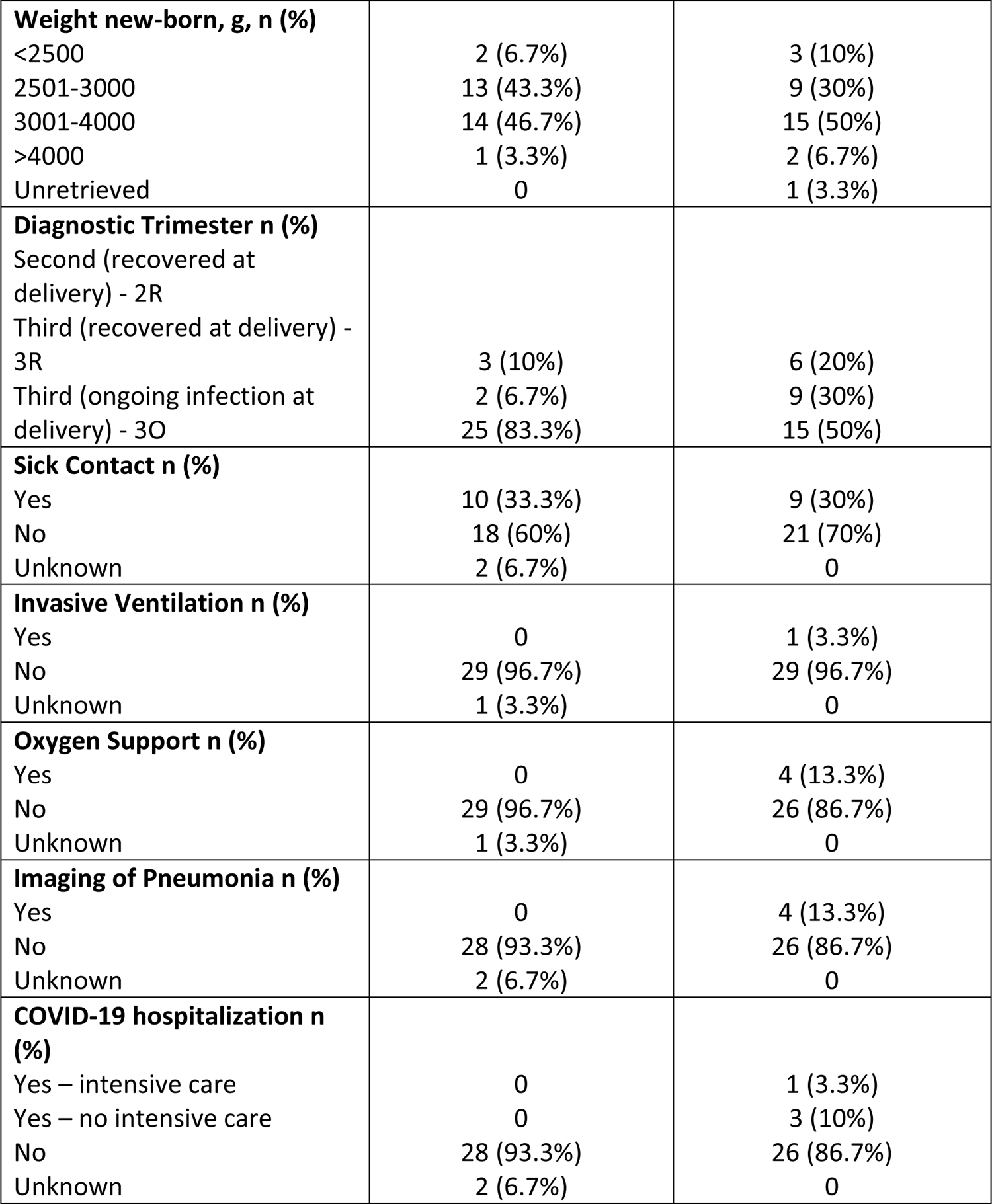
Demographic and clinical data of pregnant women infected with SARS-CoV-2 (CoV-2^+^) categorized between asymptomatic and symptomatic.

**Table 2:** Demographic and clinical data of pregnant women negative for SARS-CoV-2 (CoV-2^-^).

| SARS-CoV-2 negative PCR in mothers | n=12 |
| --- | --- |
| **Age Categories n (%)** |  |
| 18-26 | 2 (16.7%) |
| 27-33 | 5 (41.7%) |
| 34-40 | 5 (41.7%) |
| **Ethnicity n (%)** |  |
| African | 3 (25%) |
| Asian | 1 (8.3%) |
| Caucasian | 7 (58.3%) |
| Unretrieved | 1 (8.3%) |
| **Delivery Mode n (%)** |  |
| Vaginal | 4 (33.3%) |
| Caesarean | 7 (58.3%) |
| Forceps | 1 (8.3%) |
| **Gender new-born n (%)** |  |
| Female | 3 (25%) |
| Male | 9 (75%) |
| **Weight new-born, g, n (%)** |  |
| <2500 | 1 (8.3%) |
| 2501-3000 | 1 (8.3%) |
| 3001-4000 | 10 (83.3%) |

**Table 3:** Demographic and clinical data of pregnant women vaccinated with mRNA COVID-19 BNT162b vaccine.

|  |  |
| --- | --- |
| **Vaccinated mothers with mRNA COVID-19 vaccine** | n=9 |
| **Age Categories n (%)** |  |
| 18-26 | 0 |
| 27-33 | 4 (44.4%) |
| 34-40 | 5 (55.6%) |
| **Ethnicity n (%)** |  |
| Caucasian | 9 (100%) |
| **Delivery Mode n (%)** |  |
| Vaginal | 2 (22.2%) |
| Caesarean | 3 (33.4%) |
| Forceps | 2 (22.2%) |
| Unretrieved | 2 (22.2%) |
| **Gender new-born n (%)** |  |
| Female | 3 (33.4%) |
| Male | 4 (44.4%) |
| Unretrieved | 2 (22.2%) |
| **Weight new-born, g, n (%)** |  |
| <2500 | 0 |
| 2501-3000 | 2 (22.2%) |
| 3001-4000 | 4 (44.4%) |
| >4000 | 1 (11.2%) |
| Unretrieved | 2 (22.2%) |

### Poor neonatal NAbs titers are associated with maternal production of IgM-NAbs and with impaired transplacental transfer of IgG-NAbs in symptomatic infections

The transfer of antibodies from maternal to fetal circulation relies on the neonatal Fc receptor (FcRn) expressed on the placenta. Crucially, FcRn are capable of transporting antibodies of the IgG isotype, but not of the IgA nor IgM isotypes (9). Hence, detection of IgA and IgM antibodies in cord blood is used as diagnostic for fetal infection (27). Out of 50 dyads, we only detected one case where both anti-spike IgA and -IgM antibodies were present in cord blood (Figures S 1 A, B). Anti-spike IgG was found in all mothers and neonates born to SARS-CoV-2^+^ mothers in the 2R and 3R groups and in 68.4% of mothers and 56.8% of neonates born to SARS-CoV-2 mothers in the 3O group (Figure S1C). In SARS-CoV-2 recovered groups 2R and 3R, anti-spike and anti-RBD levels and titres (Figures 1 B, C) remained overall comparable within mother-neonate dyads, while there was a significant decrease in anti-spike/RBD IgG in neonates born to mothers with ongoing SARS-CoV-2 infection (Figures 1 B, C; 3O). Subsequently, the transfer ratios of anti-spike and anti-RBD IgG antibodies were ∼1 in 2R and 3R groups and ∼0.5 in the 3O group (Figure 1D), with the efficiency of antibody transfer increasing with time elapsed between SARS-CoV-2 infection and delivery (Figure 1E). We then addressed whether the sex of the fetus had an impact on the immune response to infection. We did not observe any change in maternal production of anti-spike IgA, IgM or IgG antibodies in function of the sex of the fetus (Figure S1D). However, in dyads with ongoing maternal SARS-CoV-2 infection, we observed a significant decrease in anti-spike IgG and anti-RBD in male neonates (Figures 1F, S1E).

To gauge on how anti-SARS-CoV-2 antibodies operated during pregnancy, we compared the capacity of maternal and neonate’s anti-spike antibodies to neutralize viral entry. Following SARS-CoV-2 infection, only 58.6% of mothers and 10% of neonates possessed neutralizing antibodies (Figures 1 G, H), with the frequency of detection of maternal neutralizing antibodies (Figure 1I) decreasing with time elapsed since infection from ∼60% in the 3R and 3O groups to 44.4% in the 2R group, suggesting a possible waning of NAbs in infections occurring in the second trimester. Of the 50 dyads tested, we could only detect neutralizing antibodies in 5 neonates (IQR: 23-40; Figure 1J; CoV-2^+^). In contrast, in vaccinated dyads, neutralizing antibodies were present in all neonatal blood samples (Figures 1 G, H) and their titers were similar within each dyad (Figure 1J; Vac). Infected and vaccinated mothers had equivalent neutralization titers (Figure 1J), making it unlikely that the almost absence of NAbs in neonates was primarily due to failure of the infection to induce NAbs. Conversely to what we had found for anti-spike IgG transplacental transfer (Figure 1E), the transfer ratio of NAbs did not increase with time elapsed since infection (Figure S1F). We next explored how disease presentation and the nature of maternal antibody response affected NAbs transplacental transfer. First, as the RBD domain is particularly enriched in neutralizing epitopes (11–13), we used anti-RBD antibodies as surrogates to get an insight of how disease presentation would affect NAbs transplacental transfer. Presence of symptoms did not impact the maternal levels of anti-RBD nor of NAbs, including in study participants with ongoing infection (Figures S1 G, H; 3O). Crucially, the transfer of anti-RBD antibodies, but not the ones against trimeric spike protein, plumets by ∼40% in symptomatic ongoing maternal infection (Figure 1K). Anti-spike IgM and IgA have been associated with SARS-CoV-2 neutralization in both COVID-19 disease and vaccination (14–16). This raises the possibility that poor neutralizing response in neonates resulted from maternal neutralizing activity being, at least partly, mediated by IgM/IgA and thus not transferrable to the neonate. To address this hypothesis, we correlated the levels of maternal anti-spike IgA, -IgG and -IgM antibody with neutralization titers in ongoing infection and recovered groups. In ongoing infection, neutralization titers correlated with all 3 anti-spike antibody isotypes (Figures 1L, S1I) with anti-spike IgG correlating stronger with anti-spike IgA than with anti-spike IgM (Figures 1M, S1J). Furthermore, in ongoing infection neutralization was stronger in mothers that displayed both anti-spike IgG and IgM antibodies, compared to cases that only presented anti-spike IgG or anti-spike IgM (Figure 1N; 3O). Intriguingly, in recovered maternal infection, neutralizing antibody titers correlated with anti-spike IgA and -IgM, but not with anti-spike IgG (Figures 1O, S1 K; 2R+3R). In recovered infections, anti-spike IgG and -IgM did not correlate, even though we detected an association between anti-spike IgG and -IgA (Figures 1P, S1L). In recovered infection the presence of anti-spike IgG antibodies alone was insufficient to enact neutralization (Figure 1Q). The absence of maternal samples with anti-spike IgM but without anti-spike IgG, made it impossible to evaluate whether successful neutralization could be achieved by IgM alone (Figure 1Q). In contrast, in COVID-19 vaccinated women, neutralization appears to be associated with anti-spike IgG, but not with anti-spike IgM antibodies (Figure 1R), which is in agreement with the effective transfer to the neonates of NAbs that we (Figure 1J; Vac) and others (28–30) have observed.

These results indicate that sparse detection of NAbs in neonates is due to combined action of symptomatic infection particularly hindering IgG-NAbs placental transfer and of maternal production of IgA-and IgM-NAbs in response to SARS-CoV-2 infection.

### Asymptomatic maternal infection is associated with increased early immune response comprised by anti-spike IgM and NK cells

To identify putative immune markers of infection outcomes, we analyzed maternal and neonate’s cellular response in asymptomatic versus symptomatic ongoing maternal infections. As expected from the short interval spanned between infection and sample collection, the maternal frequency of B cells, class-switch IgD^-^ B cells, CD4^+^ T cells, activated CD4^+^ T cells, CCR6^+^ T cells and CXCR5^+^ T cells remained equivalent across disease presentation (Figures 2 A-F, S2A). Interestingly, asymptomatic going infection gave rise to higher frequency of NK cells in maternal circulation and in the cord blood, even though in the latter case it did not reach statistical significance (Figures 2 G, H, S2A). The implication that immediate immune response might be important to curtail infection, made us look at rapidly produced IgM antibodies, and we found that anti-spike IgM levels were indeed higher in asymptomatic ongoing infections (Figure 2I). Upon interpolating our results with clinical data, we observed that, in mothers with ongoing infection, delayed umbilical cord clamping led to higher NK cell frequency in neonates (Figure 2J). Pertinently, delayed clamping did not alter neonatal frequency of B cells (Figure 2K) or CD4^+^ T cells (Figure 2L). Similarly, delayed clamping had no impact on neonatal NK cell frequency in resolved SARS-CoV-2 infections (Figure 2M). To pinpoint long-lasting cellular immune changes brought by SARS-CoV-2 infection, we compared the B and CD4^+^ T profiles in pregnant women that had been infected or vaccinated at similar gestational ages. Delivery occurred at an average of 66.8 and of 73.9 days post diagnosis in 2R+3R groups or inoculation of second vaccine dose, respectively. Upon maternal SARS-CoV-2 infection, the frequency of B and CD4^+^ T cells was equivalent between SARS-CoV-2 infected and COVID-19 vaccinated group (Figures S2 A-C). When we looked in more detail to CD4^+^ T cell activation and migratory status, we observed that SARS-CoV-2 infection did not alter CD4^+^ T cell activation state (Figure S2D) but led to an increase in the frequency of CXCR5^+^ T cells, and to a lesser extent of CCR6^+^ T cells (Figure S2 E, F). Importantly, this increase in the frequency of maternal CCR6^+^ and CXCR5^+^ T cells was not reflected in the fetus (Figures S2 G, H). CXCR5^+^ T cell expansion was proportional to the time elapsed between diagnosis and delivery, in asymptomatic, but not in symptomatic, infections (Figure S2I). Despite CXCR5^+^ T cells role in directing antibody production (31), their expansion was not associated with higher anti-spike IgG, IgA, or IgM levels (Figures S2 J-L).

**Figure 2.**
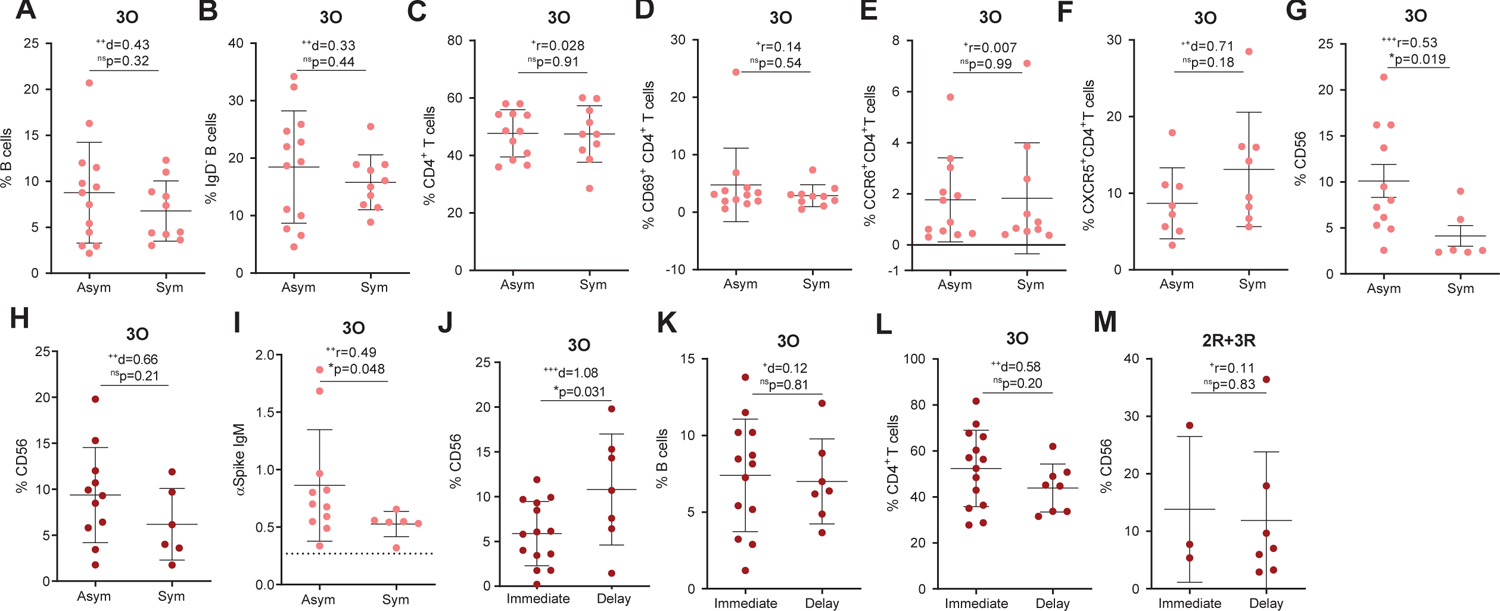
Asymptomatic maternal infection is associated with increased early immune response comprised by anti-spike IgM and NK cells. A-Maternal B cell frequency from 3O group in function of the presence (Sym) or absence (Asym) of symptoms (n = 23). B-Frequency of IgD^-^ B cells as in A. C-Frequency of CD4^+^ T cells as in A. D-Frequency of CD69^+^CD4^+^ T cells as in A. E-Frequency of CCR6^+^CD4^+^ T cell as in A. F-Frequency of CXCR5^+^CD4^+^ T cell as in A. G-Maternal NK cell frequency in participants diagnosed for SARS-CoV-2 within 7 days of delivery in asymptomatic (Asym) and symptomatic (Sym) individuals (n = 17). H-Cord blood NK cell frequency from participants diagnosed for SARS-CoV-2 within 7 days of delivery, in function of the presence (Sym) or absence (Asym) of symptoms (n = 17). I-Maternal anti-spike IgM levels as in G. J-Cord blood NK cell frequency upon either immediate or delayed umbilical cord clamping, in 3O group (n = 21). K-Cord blood B cell frequency as in J. L-Cord blood CD4^+^ T cell frequency J. M-Cord blood NK cell frequency upon either immediate or delayed umbilical cord clamping, in 2R+3R group (n = 10). p values *p < 0.05; ns, not significant were determined by unpaired t test (A, B, C, F, H J, K, L), Mann-Whitney test (D, E, G, M). Effect size measures ^+++^high, ^++^medium, ^+^small were determined by d = Cohen’s d (A, B, C, F, H, J, K, L), r = correlation coefficient r (D, E, G, M).

Altogether, our results suggest that an early immune response conveyed by anti-spike IgM antibodies and NK cells is associated with asymptomatic maternal infection and that placental transfer of NK cells might consist of a maternal sponsored mechanism to protect the fetus from infection.

### Maternal inflammatory response balances antibody placental transfer with NK cell expansion

Cytokines are key players in coordinating a healthy pregnancy (32). Nonetheless, inflammatory responses during pregnancy have been implicated in pregnancy complications (32), decreased rate of IgG transplacental transfer (9), and neurodevelopmental deficits (32). We performed a 13-cytokine multiplex assay on plasma isolated from maternal and cord blood samples. To sort out long-term from short-term cytokine alterations, we started by comparing the inflammatory profile of mothers with recovered (2R+3R) or ongoing (3O) infection. Mothers with ongoing infection exhibited higher levels of the anti-viral mediator IFN-α2, the inflammatory cytokines IL-33 and TNF-α, the anti-inflammatory cytokine IL-10, and the chemokine MCP-1 (Figure. 3A). To correct for cytokine alterations intrinsic to pregnancy and delivery (33), we compared the inflammatory profiles of participants that had been either infected or vaccinated at similar gestational age (Figure S3D). Maternal infection led to higher concentration of maternal IL-6 and IL-18, two cytokines that have been implicated in pregnancy complications and preterm delivery (32), when compared to vaccination (Figure S3D). Of the analyzed cytokines, we observed positive correlations between IL-12p40 and IFN-γ, IL-23 and IL-17, and IL-23 and IL-18 (Figures S3 A-C). While maternal serum concentration of IFN-a2 and MCP-1 inversely correlated with time elapsed between infection and delivery (Figure 3B), reinforcing their role in acute infection; elevation of inflammatory IL-6 and IL-18 was independent of time of infection (Figure. 3B). These data suggest that SARS-CoV-2 infection can lead to durable inflammatory responses long-after the infection has been cleared.

**Figure 3.**
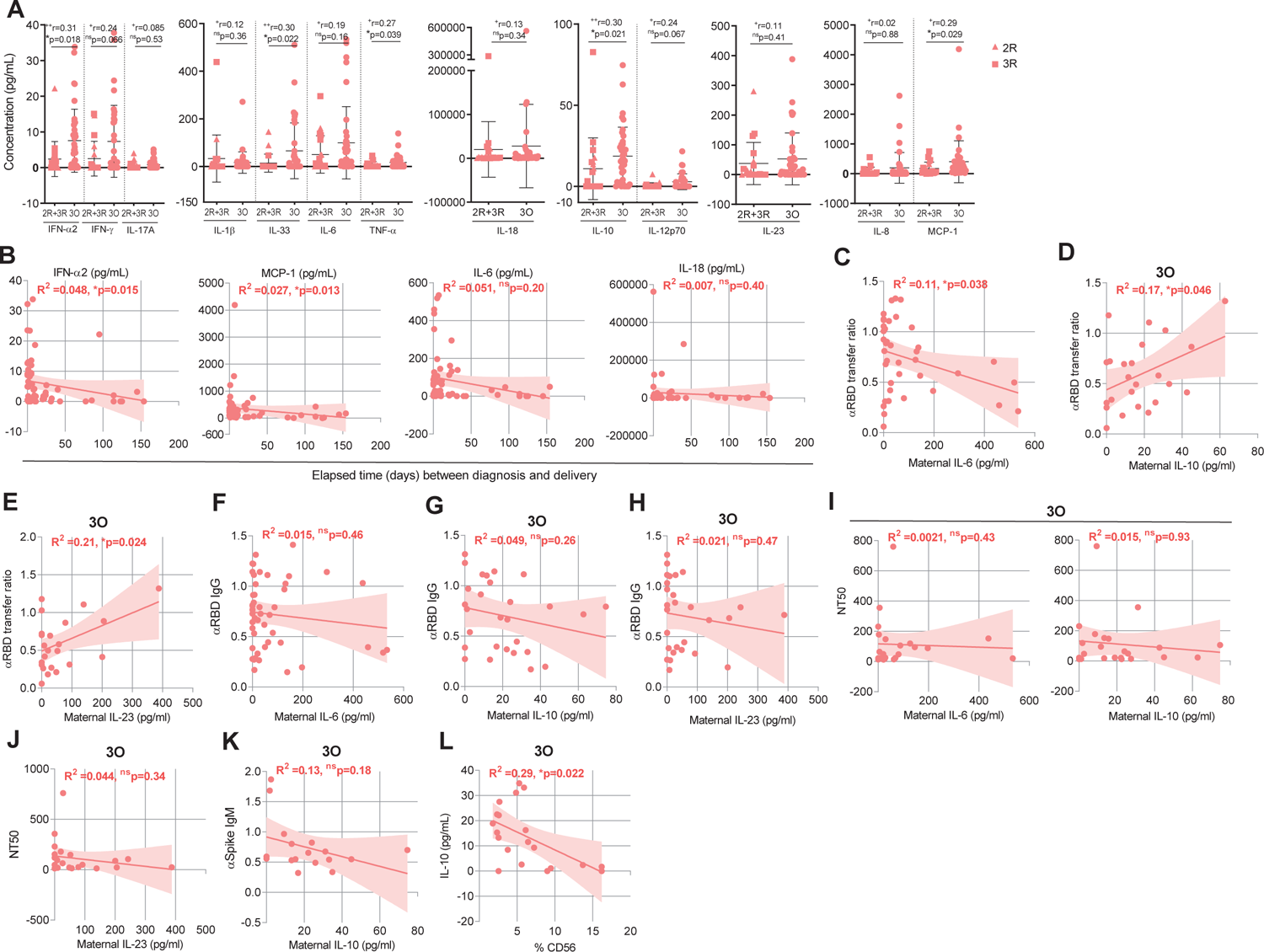
Maternal inflammatory response balances antibody placental transfer with NK cell expansion. A-Maternal plasma concentration (ng/mL) of IFN-α2, IFN-γ, IL-17A, IL-1β, IL-33, IL-6, TNF-α, IL-18, IL-10, IL-12p70, IL-23, IL-10 and MCP-1 in recovered (2R+3R, n = 20) and ongoing (3O, n = 38) infections. B-Correlation between maternal concentration of IFN-α2, MCP-1, IL-6, IL-18 and elapsed time between diagnosis and delivery (n = 58). C-Correlation between anti-RBD IgG transfer ratio and IL-6 concentration in maternal plasma (n = 39). D-Correlation between anti-RBD IgG transfer ratio and IL-10 concentration in maternal plasma in the 3O group (n = 24). E-Correlation between anti-RBD IgG transfer ratio and IL-23 concentration in maternal plasma in the 3O group (n = 24). F-Correlation between anti-RBD IgG levels and IL-6 concentration in maternal plasma (n = 48). G-Correlation between anti-RBD IgG levels and IL-10 concentration in maternal plasma in the 3O group (n = 28). H-Correlation between anti-RBD IgG levels and IL-23 concentration in maternal plasma in the 3O group (n = 28). I-Correlation between NT50 and IL-6 (left) or IL-10 (right) concentration in maternal plasma in the 3O group (n = 19). J-Correlation between NT50 and IL-23 concentration in maternal plasma in the 3O group (n = 19). K-Correlation between anti-spike IgM levels and IL-10 concentration in maternal plasma within 7 days between diagnosis and delivery (n = 17). L-Correlation between IL-10 concentration and NK cell frequency in 3O group (n = 36). p values *p < 0.05; ns, not significant were determined by Mann-Whitney test (A), Spearman correlation (B, F, I, K, L) and Pearson correlation (C, D, E, G, H). Effect size measures ^+++^high, ^++^medium, ^+^small were determined by r = correlation coefficient r (A).

Next, we sought to probe how the inflammatory response imparted on anti-RBD IgG transplacental transfer in ongoing infections (Figure 1L). We found that anti-RBD IgG transplacental transfer inversely correlated with IL-6 (Figure 3C), but positively correlated with IL-10 and with IL-23 (Figures 3 D, E). No association was detected between IL-6, IL-10, and IL-23 concentration and anti-RBD IgG or NT50 levels in maternal plasma (Figures 3 F-J).

Lastly, we sought to scrutinize how maternal inflammatory response would impact on the early NK cell and IgM immune response in asymptomatic infection (Figures 2 G-I). Curiously, IL-10 inversely associated with NK cell frequency, and no association was found between IL-10 and anti-spike IgM secretion (Figures 3 K, L). Anti-spike IgM levels and NK cell frequency were not associated with either IL-6 or IL-23 (Figures S3 E-H).

Altogether, these data indicate that IL-6 and IL-10 mediate a tug-of-war between asymptomatic viral clearance and transfer of protective anti-RBD IgG to the neonate.

### Higher concentration of at least one inflammatory cytokine in the cord blood in half of mother:neonate dyads

Even mild maternal immune responses to viral infections have resulted in adverse long-term outcomes associated to fetal inflammatory cytokines exposure (32). Of our 50 mother:neonate dyads, 27 neonates displayed higher concentration of at least one inflammatory cytokine, apart from IL-33 which was not elevated in any of the cord blood samples (Figures 4 A, B). Interestingly, polyfunctional cytokine response appears to be enriched in the cord blood of resolved infections (Figures 4B; 2R), highlighting a possible impact of gestational age in long-term neonatal outcomes. We next evaluated the contribution of maternal symptomatology and neonatal sex to the increased inflammatory profile of cord blood. In dyads with cord/maternal ratio higher than 1.5 for at least one cytokine, there was a trend for increased IFN-α2, IL-10, IL-23 and TNF-α in neonates born to symptomatic mothers (Figure 4C). When segregated by neonate’s sex, IFN-α2, IL-10 and TNF-α appear to be enriched in female neonates while IL-8 and IL-18 appear to be increased in male neonates (Figure 4D).

**Figure 4.**
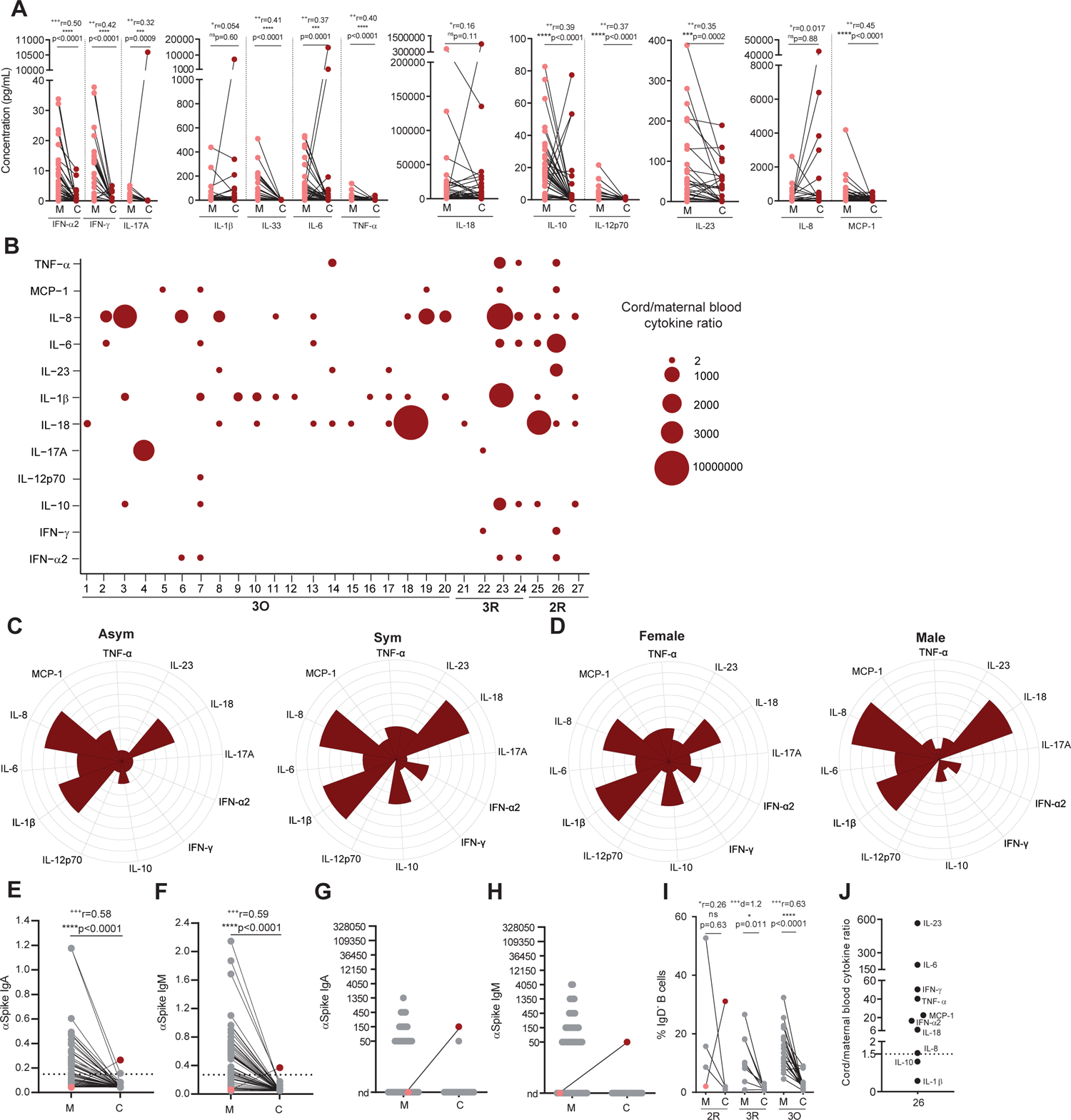
Higher concentration of at least one inflammatory cytokine in the cord blood in half of mother:neonate dyads. A-Plasma concentration (ng/mL) of IFN-α2, IFN-γ, IL-17A, IL-1β, IL-33, IL-6, TNF-α, IL-18, IL-10, IL-12p70, IL-23, IL-10 and MCP-1 in paired mother: neonate dyads (n = 50 dyads). B-Cytokine ratio between cord and maternal blood (n=27 dyads). C-Polar plots depicting the cytokine profile of neonates born to asymptomatic (left) and symptomatic (right) mothers, in dyads with cord/maternal ratio>1.5 (n=27 dyads). D-Polar plots depicting the cytokine profile of female (left) and male (right) neonates, in dyads with cord/maternal ratio>1.5 (n=27 dyads). E-Paired dyad analysis of anti-spike IgA levels. Dashed line indicates assay cutoff (n = 50 dyads). F-Paired dyad analysis of anti-spike IgM levels. Dashed line indicated assay cutoff (n = 50 dyads). G-Endpoint titers for anti-spike IgA measured in paired dyads; nd: not detectable (n = 50 dyads). H-Endpoint titers for anti-spike IgM measured in paired dyads; nd: not detectable (n = 50 dyads). I-Frequency of IgD^-^ B cells in paired dyads in 2R, 3R and 3O groups (n=30 dyads). J-Cytokine ratio between cord and maternal blood. p values *p < 0.05, ***p <0.001, ****p < 0.0001; ns, not significant were determined by non-parametric paired Wilcoxon test (A, E, F, I) and parametric paired t test (I). Effect size measures ^+++^high, ^++^medium, ^+^small were determined by r = correlation coefficient r (A, E, F, I) and d = Cohen’s d (I).

SARS-CoV-2 vertical transmission is a rare event occurring in 0-7.7% of pregnancies (7, 34, 35). Detection of IgM, and to a lesser extent of IgA, in the cord blood represents an immune fetal response to viral infection and thus is used to document vertical transmission events (9, 27). We detected anti-spike IgA and IgM in one (out of 50) mother:neonate dyads (Figures 4 E-H) in which maternal infection had occurred during the second trimester of gestation, corresponding to a period of higher ACE-2 expression (36). In agreement with the infection timeline, maternal blood was negative for anti-spike IgA and IgM antibodies, which ruled out cross-contamination (Figures 4 E-H). Even though we cannot ascertain their antigenic specificity, IgD^-^ B cells were expanded in this cord blood (Figure 4I), further supporting fetal infection. Moreover, in this dyad all but two cytokines were present at a higher concentration in the cord blood than in the mother, conducive of a fetal inflammatory response.

All in all, these data underscore that even asymptomatic maternal infections elicit fetal exposure to polyfunctional inflammation, in at least 50% of infections.

## Discussion

In this study, we investigated the cellular and molecular underpinnings regulating the balance between maternal SARS-CoV-2 clearance and transplacental transfer of protection, over the course of pregnancy. Moreover, we show that while anti-viral cytokine production is short-term, SARS-CoV-2 gestational infection leads to high concentration of inflammatory cytokines can be sustained for months. Finally, we describe that even asymptomatic maternal SARS-CoV-2 infection elicits fetal exposure to a polyfunctional inflammation, in a significant proportion of neonates.

In our cohort of 81 pregnant women ∼40% were non-caucasian, affording high robustness to the results obtained as immune responses can differ in distinct population groups due to host genetic factors, differences in the prevalence of comorbidities, and/or differences in the efficiency of transplacental antibody transfer to the fetus (37). We are aware that the vaccinated group is ethnically more homogeneous than the SARS-CoV-2^+^ cohort. Even though this disparity is bound to be multifactorial, it mainly reflects socio-economic determinants in the allocation of essential workers - at a time when vaccines were not yet available to the general population- and the ethnic diversity of catchment area of the maternity enrolling SARS-CoV-2^+^ participants. In our cohort, 3 neonates tested positive for SARS-CoV-2 by PCR. However, we could not exclude that these infections occurred post-nataly, rather than in utero. Indeed, umbilical cord testing for anti-spike IgM, which more accurately identifies in utero infections (27, 38) was negative in all these 3 cases. Distinct from recent studies (7, 8, 36, 39), our SARS-CoV-2^+^ cohort encompassed infections in the second trimester, earlier third trimester and ongoing infections. This allowed us to compare in a gestational age matched manner humoral, cellular and inflammatory immune responses following SARS-CoV-2 infection and COVID-19 vaccination.

Vertical transfer of SARS-CoV-2 antibodies, either in utero or via breastmilk, is purported to provide protection to the neonate over the first few months of life (3, 7, 8, 40–44). In SARS-CoV-2 maternal infection, altered glycosylation has been proposed to account for impaired transplacental transfer of anti-spike IgG (45). We show that the time window of decreased anti-spike IgG transfer is relatively short (∼11 days) and, as previously reported (39), affected more pronouncedly IgG transfer to male fetus. Indeed, second and earlier third trimester infections displayed anti-spike IgG transfer ratios of ∼1, in the range of what has been observed for other acute viral infections (46–48). Whether this regularization of antibody transfer with time elapsed since infection is due to a reversion of anti-spike IgG glycosylation patterns (45) once inflammation is subdued, or a result of an extended window for antibody transfer to occur, or a combination of the two still needs to be addressed. Consistent with previous works (28–30), we observed that maternal SARS-CoV-2 infection elicited similar NAbs titers to mRNA vaccination. Nonetheless, we noticed a decrease in the frequency of NAbs detection in second trimester infections. This might be due to NAbs waning, to a decrease in NAbs production in the second, and more tolerogenic (32), trimester of pregnancy, or to a combination of the two.

Distinctively from vaccination, SARS-CoV-2 maternal infection led to an almost absence of NAbs in cord blood, independently of gestational age at time of infection. Reduced transplacental transfer NAbs upon maternal SARS-CoV-2 infection had been previously noted (49–51) but the factors that govern it were yet to be identified. We advanced on this by identifying how both maternal inflammation and the nature of the neutralizing antibody response impinges on NAbs placental transfer.

To address the role of maternal inflammation on NAbs placental transfer, we started by circumventing the limitation of having a very low number of cord blood samples with NAbs (10%) by using anti-RBD IgG as a proxy for NAbs (11–13). Our results show that symptomatic infection is associated with reduced transplacental transfer of anti-RBD IgG. Mechanistically, we found that elevated IL-6 was associated with lower anti-RBD IgG transfer, while elevated IL-10 and IL-23 correlated with increased anti-RBD IgG transplacental transfer. IL-6, IL-10 and IL-23 could affect NAb transfer efficiency by altering NAb glycosylation and or FcRn-NAb binding avidity (9, 52). In contrast to our and others (49–51) findings, effective NAbs have been found in the cord blood after maternal SARS-CoV-2 infection (30). As the transfer efficiency of anti-RBD IgG relied on infection symptomatology, it is possible that the differences observed were due to distinct clinical profiles in the recruited cohorts.

Next, we identified how the nature of the neutralizing antibody response would impinge on NAbs placental transfer, as contrary to IgG, IgA and, particularly, IgM are not amenable to FcRn-mediated transplacental transfer (10). We found that neutralizing activity was not restricted to IgG isotype. In fact, in resolved infections neutralizing activity correlated exclusively with anti-spike IgM and IgA. In contrast, in vaccinated individuals neutralizing activity tended to be solely associated with anti-spike IgG, which might explain the efficient transplacental transfer of NAbs upon vaccination (28–30). Even though this is, to the best of our knowledge, the first report showing the dominance of IgM neutralizing antibodies in maternal SARS-CoV-2 infection, previous reports support our findings. In COVID-19 recovered individuals neutralizing activity has been shown to correlate better with anti-spike IgM than with anti-spike IgG (14–16). Furthermore, depleting COVID-19 convalescent plasma from IgM or IgG resulted in a 5.5- and 4.5-fold decrease in NT50 (17) and purified IgM and IgG fractions displayed similar neutralizing activities in an in vitro neutralization assay (16). Finally, the role of IgM antibodies in SARS-CoV-2 neutralization is also supported by the fact that NAbs with little or no somatic hypermutation potently neutralize the virus, indicating that extensive B cell maturation and isotype switching is not required for NAb development (12, 53). In previous studies, while IgM was a major contributor to neutralization, IgG also played an important role including in recovered individuals (14–17). Distinctively, we could only pinpoint a role for IgG neutralizing activity in ongoing, but not in resolved infections. This seemingly more relevant role of IgM for neutralization in maternal infections might be due to the immune alterations subjacent to pregnancy (20), which might have evolutionary evolved to balance rapid resolution of maternal infection with vertical transfer of protection to the developing fetus.

NK cells and IgM antibodies play a fundamental role in providing fast protective immunity against viral infections, including SARS-CoV-2 (20, 54). We uncovered an early immune response conveyed by anti-spike IgM antibodies and NK cells, which is associated with asymptomatic maternal infection, raising the possibility that NK cells might play an important role not only in controlling the development of severe COVID-19 (19), but also in governing the transition from infection to disease. Mechanistically, we unveiled that maternal IL-10 production inversely correlated with NK cell frequency. IL-10 has been reported to promote NK cell antiviral response during acute viral infection (55). Thus, it is possible that the decrease in circulating NK cell frequency is the result of IL-10 mediated activation and recruitment to the infection site. When we interpolated our results with clinical data, we remarked that in mothers with ongoing infection, delayed umbilical cord clamping led to a selective enrichment of NK cells, but not of B or CD4^+^ T cells, in the cord blood. Remarkably, this enrichment in NK cell transfer to the neonate at the time of birth did not occur in recovered infections. Even though the role of maternal transferred immune system in protection the neonate protection against infection remains incompletely understood (56), these data raise the enticing possibility that placental NK cell transfer might consist of a maternal sponsored mechanism to protect the neonate, at a time of elevated immune fragility, from post-natal exposure to the infected mother. The physiological relevance of such peri-natal vertical transfer of protection via NK cells might be reinforced by the fact that it has been documented that IgG antibodies that activate NK cells are preferentially transported across the placenta (57).

Consistent with previous reports (7, 8), we show that upon SARS-CoV-2 infection, pregnant women mount an inflammatory response composed by IFN-α, IL-6, IL-10, IL-18, IL-33 and MCP-1. Intriguingly, we found that while cytokines related to acute viral response returned to normal levels upon infection clearance, the inflammatory response mediated by IL-6 and IL-18 remained elevated for weeks to months past infection resolution. Sustained inflammatory activation after asymptomatic or mild infections might bring some concerns in the future neurological development of the neonate, as maternal inflammation, including by IL-6 and IL-18, have been linked to altered immune responses (58), immune mediated diseases (59), and an increased risk of psychiatric disorders later in life (60, 61). This might be even more worrying, since we observed higher concentration of at least one inflammatory cytokine in the cord blood in ∼50% of dyads. Ex vivo studies of term placentas indicate that transplacental transfer of most cytokines does not occur (62). However, other studies indicate that maternal inflammatory responses might lead to placental cytokine production (25, 26) and transfer of maternal cytokines has been documented in in vivo animal models (63). Thus, it is possible the cytokines increased by thousand-fold magnitude in the cord blood to be neonatally derived.

A combination of uncommon SARS-CoV-2 viraemia in pregnant women (64), and negligible placental co-expression of its canonical cell entry receptors ACE2 and TMPRSS2 (65) account for the low frequency of SARS-CoV-2 vertical transmission. As FcRn selectively transports IgG across the placenta, most accurate way to identify vertical transmission is through the detection of anti-SARS-CoV-2 IgA and IgM antibodies in cord blood. Out of 50 paired mother:neonate dyads, we detected anti-spike IgA and IgM in one cord blood sample, which is consistent with previous studies that reported anti-spike IgM in 0-7.7% of cord blood samples (7, 34, 66). Curiously, our putative case of vertical transmission occurred during the second trimester, a gestational period when ACE2 placental expression peaks (36). Even though maternal infection had long resolved by the time of delivery, the cord blood displayed higher concentration of 8 distinct cytokines than the mother. Moreover, this cord blood also exhibited an exceptionally high IgD^-^ B cell population documenting a fetal adaptive immune response, likely to SARS-CoV-2 infection.

Our data point to an evolutionary trade-off which might benefit the neonate in two ways, first by skewing maternal immune response towards immediate viral clearance, and second by coinciding the optimal vertical transfer of protection, via NK cells and IgA/IgM-NAbs, to the most fragile perinatal period. Considering the energetic costs of mounting an immune response (67), which in vertebrates are equivalent to the ones of reproduction and growth (68), it might be advantageous for both the pregnant individual and fetus to concentrate the immune efforts in clearing the infection instead of splitting it between mother and fetus. First, it reasons that by rapidly clearing the viral infection, the maternal immune response effectively prevents it from reaching the developing fetus; second, local immune and placental cells provide potent protective mechanisms that effectively protect the fetus from infection(69–71). Conceivably, the priming of IgA- and IgM-NAbs together with NK cells might be best suited to convey immune protection to the neonate soon after birth. Pertinently, IgA- and IgM-NAbs producing plasma cells primed during pregnancy are known to migrate to the breast and secrete protective IgA- and IgM-NAbs to the milk (1, 3, 40, 72). We showed that NK cells are selectively transferred perinatally in ongoing maternal infections. Thus, gestational IgM and NK cell priming and perinatal transfer might consist in a seemingly maternal sponsored mechanism of protection to the neonate, at a time when there is an increased risk of horizontal viral transmission, due to close contact with the mother, and higher fragility of the newborn, being exposed for the first time to the outside environment without the placental protective barrier.

## Methods

### Biospecimen collection

A total of 79 peripheral and 69 cord blood samples were collected at the time of delivery from 60 SARS-CoV-2 infected pregnant women, 12 non-infected pregnant controls, and 9 from COVID-19 mRNA vaccinated pregnant women. SARS-CoV-2 positive and negative participants were recruited between May 2020 and February 2021, before vaccination had become available to the general population. Vaccinated participants were recruited between July 2021 and February 2022. Nasopharyngeal swabs were obtained for all study participants upon hospital admission. Nasopharyngeal swabs of the babies were collected whenever possible. All women underwent clinical evaluation of vital signs and symptoms, laboratory analysis and radiological chest assessment at the discretion of physicians. Therapeutic management was consequently tailored according to clinical findings and national guidelines. Demographic and clinical characteristics are detailed in Tables 1, 2 and 3. Cord blood was obtained through venipuncture of the cord vein. Blood samples were immediately processed.

### Exclusion criteria

In total, we recruited 63 pregnant women suspected of SARS-CoV-2 infection. Three participants were excluded from the study due to loss of biospecimens’ integrity. One participant was excluded from the control group due to prior exposure to SARS-CoV-2.

### Peripheral and cord blood mononuclear cells (PBMCs and CBMCs) isolation

Peripheral and cord blood samples were collected in EDTA tubes. PBMCs and CBMCs were isolated by density gradient centrifugation (Biocoll, Merck Millipore) (3, 73), cryopreserved in 10% DMSO in FBS and stored at −80°C until subsequent analysis. Plasma samples were carefully removed from the cellular fraction and stored at −80°C, until further analysis.

### ELISA

Antibody binding to SARS-CoV-2 trimeric spike protein or its RBD domain was assessed by a previously described in-house ELISA assay (74) based on the protocol by Stadlbauer *et al* (75). Briefly, 96-well plates (Nunc) were coated overnight at 4°C with 0.5 mg/ml of trimeric spike or RBD. After blocking with 3% BSA diluted in 0.05% PBS-T, 1:50 diluted plasma was added and incubated for 1 h at room temperature. Plates were washed and incubated for 30 min at room temperature with 1:25,000 dilution in 1% BSA 0.05% PBS-T of HRP-conjugated anti-human IgA, IgG and IgM antibodies (Abcam, ab97225/ab97215/ab97205). Plates were washed and incubated with TMB substrate (BioLegend), stopped by adding phosphoric acid (Sigma) and read at 450nm. Cut-off for plasma samples resulted from the mean of OD_450_ values from negative controls plus 3 times the standard deviation (74). Endpoint titers were established using a 3-fold dilution series starting at 1:50 and ending at 1:109,350 and defined as the last dilution before the signal dropped below OD_450_ of 0.15. This value was established using plasma from pre-pandemic samples collected from subjects not exposed to SARS-CoV-2 (74). For samples that exceeded an OD_450_ of 0.15 at last dilution (1:109,350), end-point titter was determined by interpolation (76). As previously described (74), in each assay we used 6 internal calibrators from 2 high-, 2 medium- and 2 low-antibody producers that had been diagnosed for COVID-19 through RT-PCR of nasopharyngeal and/or oropharyngeal swabs. As negative controls, we used pre-pandemic plasma samples collected prior to July 2019.

### Production of ACE2 expressing 293T cells

Production of 293T cells stably expressing human ACE2 receptor was done as previously described (77). Briefly, for production of VSV-G pseudotyped lentiviruses encoding human ACE2, 293T cells were transfected with pVSV-G, psPAX2 and pLEX-ACE2 using jetPRIME (Polyplus), according to manufacturer’s instructions. Lentiviral particles in the supernatant were collected after 3 days and were used to transduce 293T cells. Three days after transduction, puromycin (Merck, 540411) was added to the medium, to a final concentration of 2.5 μg/ml, to select for infected cells. Puromycin selection was maintained until all cells in the control plate died and then reduced to half. The 293T-Ace2 cell line was passaged six times before use and kept in culture medium supplemented with 1.25 μg/ml puromycin.

### Production of spike pseudotyped lentivirus

To generate spike pseudotyped lentiviral particles, 6×10^6^ 293ET cells were co-transfected with 8.89 mg pLex-GFP reporter, 6.67 mg psPAX2, and 4.44 mg pCAGGS-SARS-CoV-2-S_trunc_ D614G, using jetPRIME according to manufacturer’s instructions. The virus-containing supernatant was collected after 3 days, concentrated 10 to 20-fold using Lenti-XTM Concentrator (Takara, 631231), aliquoted and stored at −80°C. Pseudovirus stocks were titrated by serial dilution and transduction of 293T-Ace2 cells. At 24h post transduction, the percentage of GFP positive cells was determined by flow cytometry, and the number of transduction units per mL was calculated.

### Neutralization assay

Heat-inactivated plasma was four-fold serially diluted and then incubated with spike pseudotyped lentiviral particles for 1h at 37°C. The mix was added to a pre-seeded plate of 293T-Ace2 cells, with a final MOI of 0.2. At 48h post-transduction, the fluorescent signal was measured using the GloMax Explorer System (Promega). The relative fluorescence units were normalized to those derived from the virus control wells (cells infected in the absence of plasma), after subtraction of the background in the control groups with cells only.

### Immune phenotyping of maternal and cord blood leukocytes

For immunophenotyping of B, T and NK cells, cryopreserved PBMCs and CBMCs were rested for 1h at 37°C and stained with a fixable viability dye eFluor^TM^ 506 (Invitrogen) and surface labelled with the following antibodies all from BioLegend: anti-CD3 (300424/UCHT1), anti-CD4 (344666/SK3), anti-CD69 (310910/FN50), anti-CXCR5 (356904/J252D4), anti-CCR6 (353432/G034E3), anti-CD19 (363026/SJ25C1), anti-IgD (348250/IA6-2), and anti-CD56 (318348/HCD56). Cells were washed, fixed with 1% PFA and acquired in BD FACS Aria III (BD Biosciences) and analyzed with FlowJo v10.7.3 software (Tree Star).

### Luminex

Plasma samples were thawed and tested in the 13-plex LegendPlex Human Inflammation panel 1 (BioLegend, 740809) to quantify levels of IL-1β, IFN-α2, IFN-γ, TNF-α, MCP-1, IL-6, IL-8, IL-10, IL-12p70, IL-17A, IL-18, IL-23 and IL-33. The assay was performed according to manufactor’s instructions and was modified by using half of the amount of all reagents. All plasma samples were diluted 2X with assay buffer, and sample concentrations were calculated according to the dilution factor. Briefly, 12.5 µl of diluted plasma or standard, and 12.5 µl of mixed beads were added to each well and incubated for 2 hours. The plate (V-bottom 96 well plate) was washed twice with 100 µl of wash buffer. Samples and standards were incubated with 12.5 µl of detection antibody for 1 hour followed by an incubation of 30 minutes with 12.5 µl of Streptavidin-PE. The plate was washed once, and samples were resuspended in 75 µl of wash buffer. All incubation steps were performed at 800 rpm at room temperature and protected from the light. Samples were acquired in a BD FACS Canto (BD Biosciences) and analyzed with the Windows LegendPlex software (v8.0 BioLegend).

### Statistical Analysis

Statistical analysis was performed by using GraphPad Prism v9.00. First, we tested the normality of the data by using D’Agostino & Pearson normality test, by checking skewness and kurtosis values and visual inspection of data. Then, if the samples followed a normal distribution, we chose the appropriate parametric test; otherwise, the non-parametric counterpart was chosen. In two groups comparison: for paired data the Wilcoxon matched-pairs signed-rank test and paired t test were used; for unpaired data, Man-Whitney test and the unpaired t test were used. For multiple groups comparison, ordinary one-way analysis of variance (ANOVA) with posttest Holm-Sidàk’s multiple comparisons or Kruskal-Wallis tests with posttest Dunn’s multiple comparisons were used as indicated in figure legends. Spearman and Pearson correlation tests were used in correlation analysis as described. The ratios of cord/maternal ratio for inflammatory cytokines were imported into an excel spreadsheet and analyzed in R (v2022.2.3.0) to generate matrices with ggplot package. Polar plots were generated in Origin 2022. The half-maximal neutralization titer (NT50), defined as the reciprocal of the dilution at which infection was decreased by 50%, was determined using four-parameter nonlinear regression (least-squares regression without weighting; constraints: bottom = 0). To measure the magnitude of the difference, the effect size was calculated as described (78, 79)-for Paired t test: Cohen’s d (d) is small if <0.3, medium if ≥0.3 or large if ≥0.8; -for Wilcoxon test and Man-Whitney test: correlation coefficient r (r) is small if <0.3, medium if ≥0.3 or large if ≥0.5; -for Kruskal-Wallis and ordinary one-way ANOVA: eta square (η^2^) is small if <0.01, medium if ≥0.06 or large if ≥0.14. Effect sizes values are reported in figure legends and are labeled as ^+^ for small, ^++^ for medium and ^+++^ for large.

### Study Approval

All participants provided informed consent and all procedures were approved by the ethics committees of Centro Hospitalar de Lisboa Central (859/2020) and of NOVA Medical School (112/2021/CEFCM), in accordance with the provisions of the Declaration of Helsinki and the Good Clinical Practice guidelines of the International Conference on Harmonization.

## Author contributions

J.G., M.M., M.A., and J. C.-N. designed and performed experiments and analyzed the data. N.C., F.S., J.G., and H.S. enrolled the subjects and collected demographic data. M.J.A., C.A., and C.P. provided critical expertise. H.S. conceptualized the study, designed experiments, analyzed the data, supervised the project, and wrote the manuscript. All authors discussed the results and commented on the manuscript.

## Data Availability

All data produced in the present study are available upon reasonable request to the authors

## Acknowledgements

We are very grateful to all study participants and to all the CHULC doctors and nurses that contributed to this study. We thank Cláudia Andrade (Flow Cytometry Platform), Daniela Amaral-Silva, Manuela Correia, Ricardo Santos and Margarida Archer for support. This work was funded by European Society of Clinical Microbiology and Infectious Diseases (ESCMID) and by Gilead Génese (PGG/009/2017) grants to H.S., RESEARCH4COVID 19 (Ref 580) to M.J.A. M.J.A., H.S. and J.G are supported by FCT through work contracts CEECIND/02373/2020, CEECIND/01049/2020 and PTDC/MEC-REU/29520/2017, respectively. Graphical abstract was created using BioRender.com.

---

**Figure S1.**
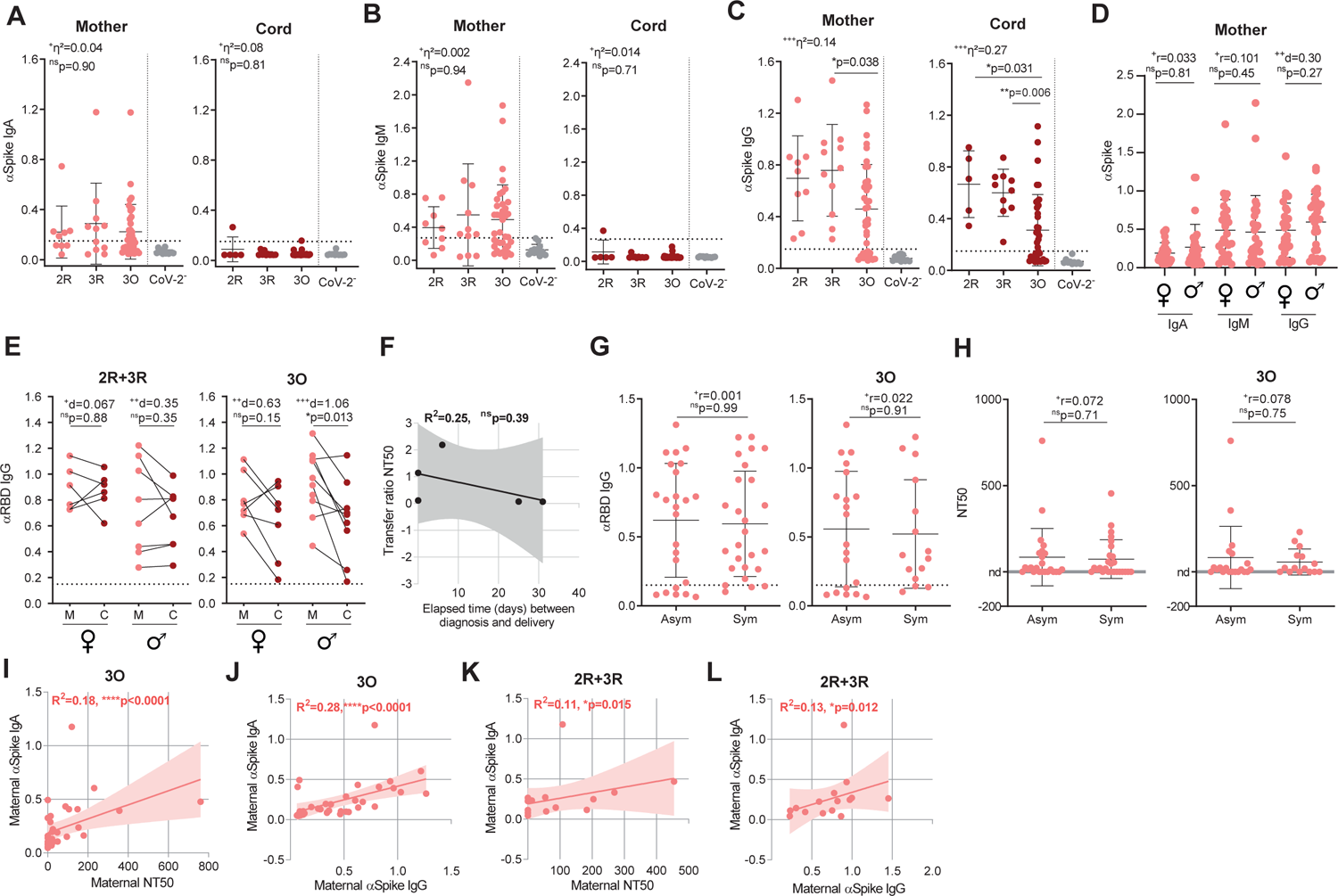
Antibody levels in maternal:neonate dyads classified by sex of the neonate and symptomatology. A-Anti-spike IgA levels in maternal and cord blood samples of SARS-CoV-2 positive mothers divided by 2R, 3R and 3O groups and in non-infected maternal/cord controls (CoV-2^-^, grey), measured by absorbance at 450 nm (OD_450_). Dashed line indicates assay cutoff (CoV-2^+^ n = 60; CoV-2^-^ n = 12). B-Anti-spike IgM levels as in A. C-Anti-spike IgG levels as in A. D-Anti-spike IgA, IgM and IgG levels in plasma of SARS-CoV-2 positive mothers according to the neonate sex (n = 56). E-Paired dyads analysis of anti-RBD IgG in 2R+3R (left; n = 15 dyads) and 3O (right, n = 18 dyads) groups according to neonate sex. F-Correlation between NT50 transfer ratio and elapsed time between diagnosis and delivery (n = 5 dyads). G-Anti-RBD IgG levels in asymptomatic (Asym) and symptomatic (Sym) SARS-CoV-2^+^ infection in all participants (left; n = 50) or in 3O group only (right; n = 35). H-NT50 in asymptomatic (Asym) and symptomatic (Sym) individuals in all SARS-CoV-2^+^ mothers (left; n = 50) or in 3O group only (right; n = 35). I-Correlation between maternal NT50 and anti-spike IgA in 3O group (n = 35). J-Correlation between maternal anti-spike IgA and IgG levels in 3O group (n = 35). K-Correlation between maternal NT50 and anti-spike IgA in 2R+3R groups (n = 15). L-Correlation between maternal anti-spike IgG and IgA levels in 2R+3R groups (n = 15). p values *p < 0.05, **p < 0.01, ****p < 0.0001; ns, not significant were determined by Kruskal-Wallis test (A, B, C), unpaired t test (D), Mann-Whitney test (D, F, G) parametric paired t test (E), Pearson correlation (H) and Sperman correlation (I-L). Effect size measures ^+++^high, ^++^medium, ^+^small were determined by η^2^ = eta-squared (A, B, C) d = Cohen’s d (D, E) and r = correlation coefficient r (D, F, G).

**Figure S2.**
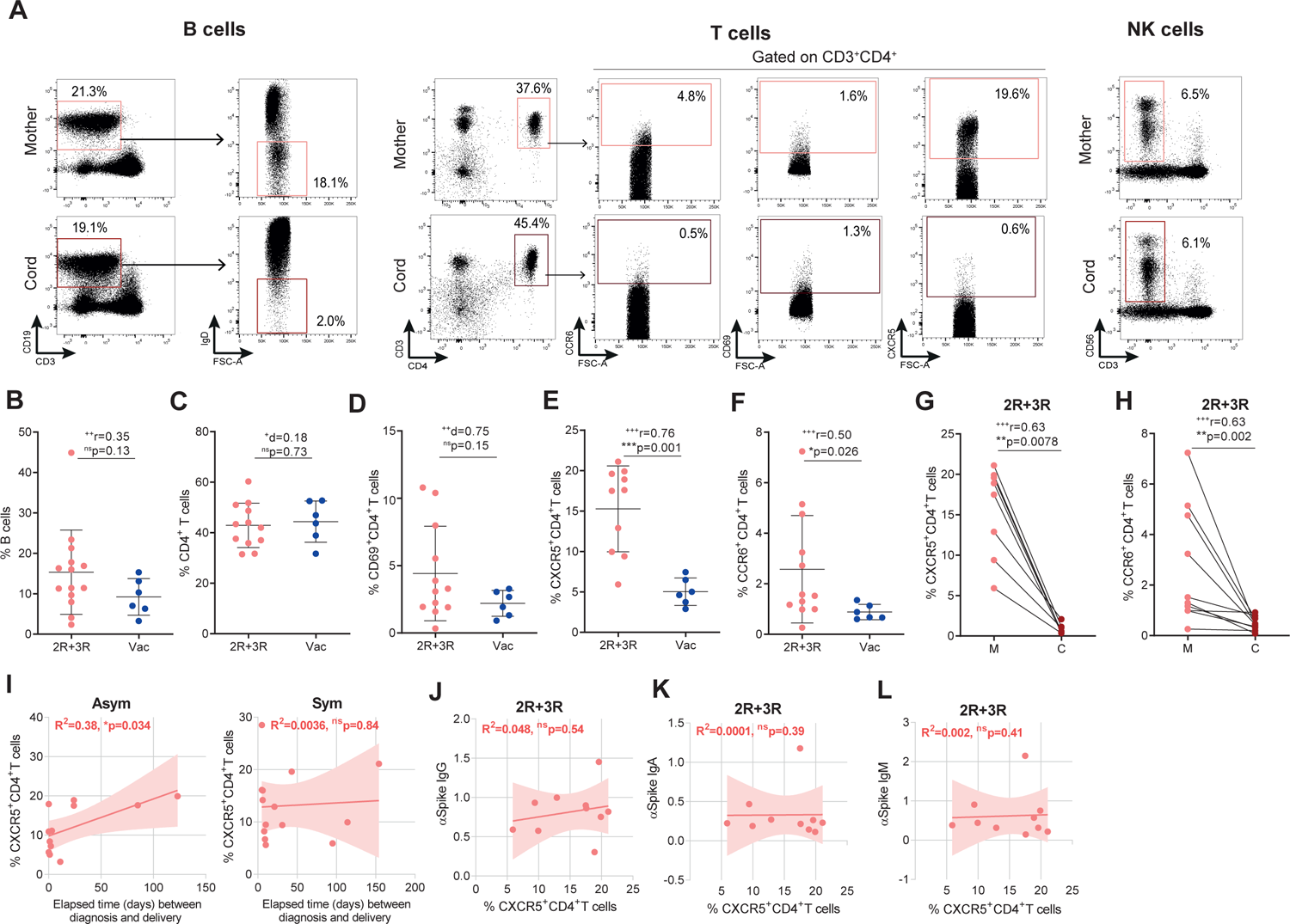
Long-term expansion of CXCR5^+^CD4^+^ T cells in maternal SARS-CoV-2 infection. A-Gating strategy for CD3^-^CD19^+^IgD^-^ B cells (left); for CD69^+^, CCR6^+^ and CXCR5^+^ CD3^+^CD4^+^ T cells (middle); and for CD3^-^CD56^+^ NK cells (right). B-Frequency of circulating B cells in SARS-CoV-2 recovered (2R+3R; n = 14) and in vaccinated participants (n = 6). C-Frequency of CD4^+^ T cells as in B. D-Frequency of CD69^+^CD4^+^ T cells as in B. E-Frequency of CXCR5^+^CD4^+^ T cells as in B. F-Frequency of CCR6^+^CD4^+^ T cells as in B. G-Paired dyad analysis of the frequency of CXCR5^+^CD4^+^ T cells in SARS-CoV-2 recovered participants (2R+3R; n = 8 dyads). H-As in G for CCR6^+^CD4^+^ T cells (n = 10 dyads). I-Correlation between the frequency of maternal CXCR5^+^CD4^+^ T cells and the elapsed time between diagnosis and delivery in asymptomatic (left) and symptomatic (right) individuals (n = 25). J-Correlation between maternal anti-spike IgG levels and the frequency of CXCR5^+^CD4^+^ T cells in 2R+3R group (n = 10). K-As in J for maternal anti-spike IgA levels. L-As in J for maternal anti-spike IgM levels. p values *p < 0.05, **p < 0.01, ***p <0.001; ns, not significant were determined by Mann-Whitney test (B, E, F), unpaired t test (C, D), non-parametric paired Wilcoxon test (G, H), Pearson correlation (I, J) and Spearman correlation (K, L). Effect size measures ^+++^high, ^++^medium, ^+^small were determined by r = correlation coefficient r (B, E, F, G, H) and d = Cohen’s d (C, D).

**Figure S3.**
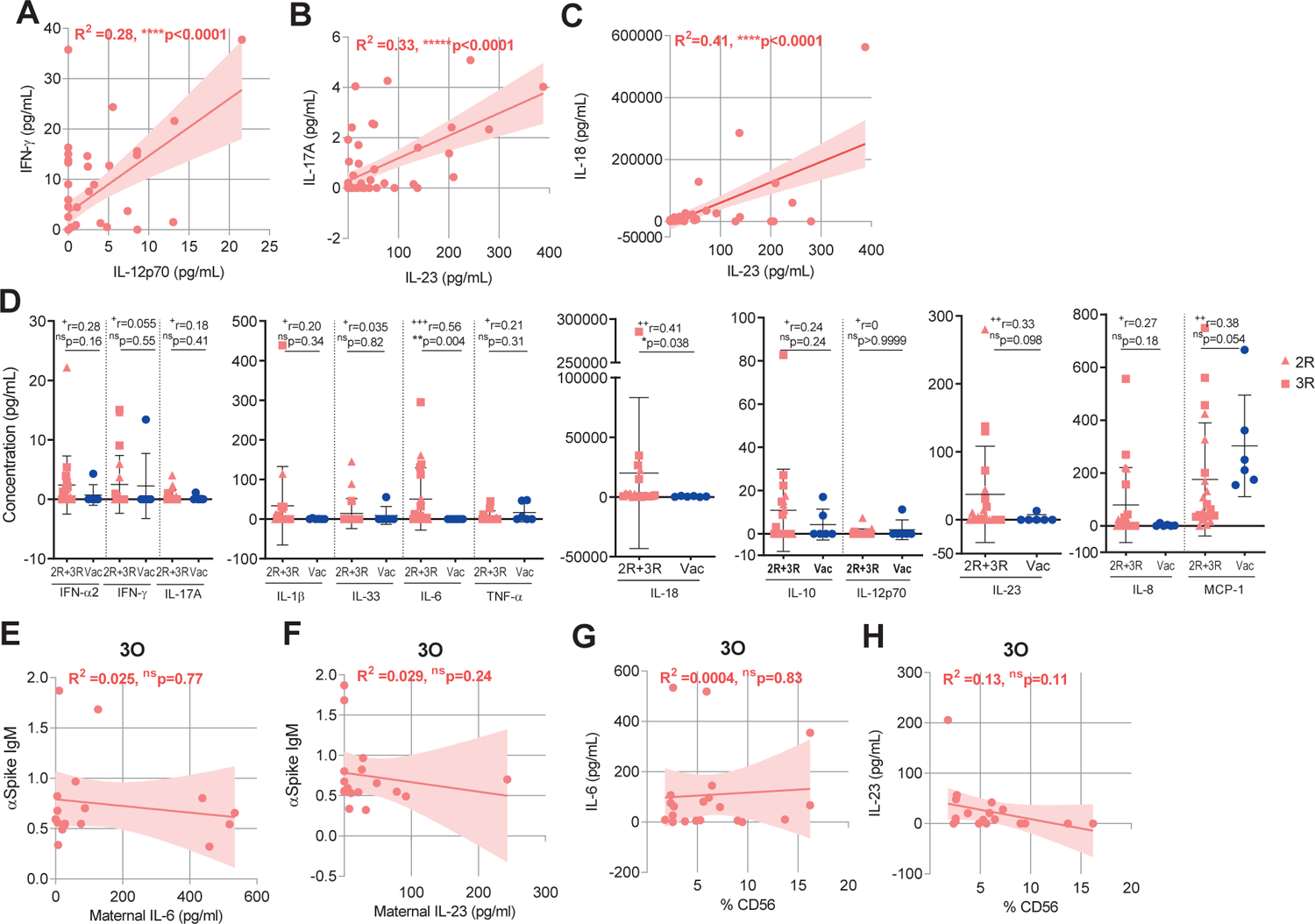
Gestational age matched inflammatory profile in SARS-CoV-2 infected and COVID-19 vaccinated individuals. A-Correlation between IFN-γ and IL-12p70 concentration in maternal plasma (n = 58). B-Correlation between IL-17A and IL-23 concentration in maternal plasma (n = 58). C-Correlation between IL-18 and IL-23 concentration in maternal plasma (n = 58). D-Plasma concentration in ng/mL of IFN-α2, IFN-γ, IL-17A, IL-1β, IL-33, IL-6, TNF-α, IL-18, IL-10, IL-12p70, IL-23, IL-10 and MCP-1 in gestational age-matched infected (CoV-2^+^ n = 20) and vaccinated (Vac n = 6) individuals. E-Correlation between maternal anti-spike IgM levels and IL-6 concentration in maternal plasma diagnosed within 7 days of delivery (n = 17). F-As in E for IL-23 concentration (n = 17). G-Correlation between maternal IL-6 concentration and NK cell frequency in 3O group (n = 36). H-As in G for IL-23 concentration. p values *p < 0.05, **p < 0.01, ****p < 0.0001; ns, not significant were determined by Spearman correlation (A-C, E-H) and Mann-Whitney test (D). Effect size measures ^+++^high, ^++^medium, ^+^small were determined by r = correlation coefficient r (D).

## Notes

### Competing Interest Statement

The authors have declared no competing interest.

### Funding Statement

This work was funded by European Society of Clinical Microbiology and Infectious Diseases (ESCMID) and by Gilead Genese (PGG/009/2017) grants to H.S., RESEARCH4COVID 19 (Ref 580) to M.J.A. M.J.A., H.S. and J.G are supported by FCT through work contracts CEECIND/02373/2020, CEECIND/01049/2020 and PTDC/MEC-REU/29520/2017, respectively.

